# The effect of smokers transitioning to E-cigarettes on physical and mental health: An emulated trial using longitudinal data

**DOI:** 10.64898/2026.02.12.26345898

**Authors:** Shaik Jasim Thameemul Ansari

## Abstract

**Introduction:** Tobacco smoking remains a leading cause of preventable death in the UK. Although e-cigarettes are promoted as a harm-reduction option, longitudinal evidence on short-term health outcomes across different smoking transition pathways is limited. This study examined short-term associations between transitions to exclusive e-cigarette use, dual use, or cessation and physical health, mental health, and health-related quality of life, compared with continued smoking.

**Methods:** A target trial emulation framework was applied to Waves 7-14 (2015-2024) of the UK Household Longitudinal Study, including 18,011 participant-wave observations from baseline smokers. Propensity score matching (1:3) was used to create comparable exposure groups. A doubly robust analysis-combining matching with Ordinary Least Squares regression-estimated outcomes using the SF-12 Physical (PCS) and Mental (MCS) Component Summary scores and a mapped EuroQol 5-Dimensions 3-Level version (EQ-5D-3L) index. The SF-12 is a validated generic health measure, where PCS and MCS are norm-based scores (mean = 50, SD = 10). The EQ-5D-3L index (range: 0 to 1) reflects overall health utility.

**Results:** Compared with continued smokers, exclusive e-cigarette users had higher short-term mental health scores (SF-12 MCS β = 1.042; 95% CI: 0.229 to 1.855). In contrast, dual users had lower mental health scores (β = -1.023; 95% CI: -1.574 to -0.472). Short-term physical health scores (SF-12 PCS) were lower among both exclusive switchers (β = -0.670; 95% CI: - 1.287 to -0.053) and quitters (β = -0.486; 95% CI: -0.853 to -0.119), with no evidence of short-term physical health improvement for any transition group. Dual users also had lower health-related quality of life (EQ-5D-3L β = -0.016; 95% CI: -0.025 to -0.008). Subgroup analyses suggested heterogeneity by age and socioeconomic position, with poorer outcomes among older and more disadvantaged smokers. Sensitivity analyses produced directionally consistent findings.

**Conclusion:** Exclusive switching to e-cigarettes was associated with higher short-term mental health scores, whereas dual use was associated with poorer mental health and health-related quality of life. These findings underscore the importance of distinguishing complete switching from dual use when designing harm-reduction policies and smoking cessation support.

## INTRODUCTION

Tobacco smoking remains one of the leading causes of preventable morbidity and mortality worldwide and continues to pose a substantial public health challenge in the UK (1). Beyond its well-established contribution to cardiovascular disease, cancer, respiratory conditions, and adverse mental health outcomes, smoking imposes a considerable economic and societal burden (2). In England alone, smoking is estimated to cost the National Health Service approximately £1.9 billion annually, with additional costs arising from lost productivity, premature mortality, smoking-related unemployment, and long-term social care needs (3, 4). In response, electronic cigarettes (e-cigarettes) have been promoted as a harm-reduction and smoking cessation tool for adult smokers. However, despite their growing use and policy relevance, the evidence base supporting these strategies remains constrained by important methodological limitations, such as fail to adjust adequately for confounding. Concerns also persist regarding potential adverse physical and mental health outcomes associated with both smoking and e-cigarette use, including respiratory and cardiovascular symptoms, neurological effects, and associations with anxiety, depression, psychological distress, and cognitive impairment (5–7). These uncertainties underscore the need for effective and evidence-based cessation and harm-reduction strategies.

Evidence to date presents a mixed picture. Randomised controlled trials and systematic reviews suggest that nicotine-containing e-cigarettes can be more effective than traditional nicotine replacement therapies in supporting smoking cessation under structured conditions, and several government and public health bodies in the UK have endorsed their use for this purpose(8, 9). However, findings from observational studies are less consistent, particularly when examining health outcomes beyond cessation itself. A key challenge is the inconsistent definition and categorisation of smoking behaviour. Many studies treat smoking status as static or fail to clearly distinguish between exclusive switching to e-cigarettes, dual use of cigarettes and e-cigarettes, and complete cessation (10, 11). This limits the ability to meaningfully interpret associations with physical health, mental health, and health-related quality of life (HRQoL), and complicates attempts to separate association from causation.

Methodological limitations are especially pronounced in observational research. Confounding by indication (12)is a major concern, as individuals who switch to e-cigarettes or quit smoking may differ systematically from those who continue smoking in terms of baseline health, motivation, or socioeconomic position (13). Follow-up periods are often short or cross-sectional, restricting the ability to capture health changes over time and increasing vulnerability to reverse causation, particularly for mental health outcomes, where deteriorating health may prompt smoking transitions rather than result from them. In addition, heterogeneity in exposure definitions across studies leads to exposure misclassification, obscuring potentially important differences between exclusive switching and dual use.

Against this backdrop, there is a clear need for robust, longitudinal evidence that is representative of the UK population and that explicitly accounts for the dynamic nature of smoking behaviour (14–18) This study addresses this gap by using nationally representative longitudinal data and clearly defined smoking transition categories, continued smokers, exclusive e-cigarette switchers, dual users, and quitters. The aim is to estimate associations between smoking transitions and physical health, mental health, and HRQoL, while strengthening causal interpretation through careful control of confounding by indication and exposure misclassification. By applying a target trial emulation framework to real-world data, this study seeks to provide timely and policy-relevant evidence to inform smoking cessation services and harm-reduction strategies in the UK.

## METHODS

### Study Design and Data Source

This study adopted a target trial emulation (TTE) framework, as proposed by Hernán and Robins (19), to strengthen causal interpretation using observational data. A longitudinal design was implemented using data from the UK Household Longitudinal Study (UKHLS), a nationally representative panel study that follows individuals and households across the UK over time and collects detailed information on health, socioeconomic circumstances, and health-related behaviours.

Analyses focused on Waves 7–14 (2015–2024), which were structured as consecutive wave pairs (e.g., Wave 7–Wave 8, Wave 8–Wave 9). Each wave pair represented a discrete emulated trial, with baseline exposure assessed at the initial wave (T0) and outcomes measured at the subsequent wave (T1).

### Eligibility Criteria

Participants were eligible if they:

- Were aged 16 years or older at baseline
- Were identified as current traditional cigarette smokers at the initial wave (T0)
- Had valid responses on smoking status and e-cigarette use across at least two consecutive waves
- Had non-missing data on health outcomes (SF-12 Physical and Mental Component Summary scores)
- Had complete data on all baseline covariates included in the analysis.

Participants who reported e-cigarette use or dual use at baseline were excluded.

### Exposure Definition: Smoking Transitions

Smoking status was assessed at baseline (T0) and follow-up (T1) to define four mutually exclusive transition groups:

- **Continued smokers**: Traditional cigarette smokers at both T0 and T1.
- **Switchers**: Traditional cigarette smokers at T0 who reported exclusive e-cigarette use at T1.
- **Dual users**: Traditional cigarette smokers at T0 who reported concurrent use of cigarettes and e-cigarettes at T1.
- **Quitters**: Traditional cigarette smokers at T0 who reported no use of cigarettes or e-cigarettes at T1.

In all analyses, continued smokers served as the reference group.

### Outcome Measures

All outcomes were measured at follow-up (T1) for each wave pair, conditioning on baseline values. Differences of approximately 3 to 5 points are generally considered clinically meaningful (20, 21):

- Physical health: SF-12 Physical Component Summary (PCS) score.
- Mental health: SF-12 Mental Component Summary (MCS) score.
- Health-related quality of life (HRQoL): EQ-5D-3L utility score, mapped from SF-12 PCS and MCS using the validated Lawrence–Fleishman algorithm (22).

### Covariates

Baseline covariates were selected a priori based on existing epidemiological evidence and causal assumptions regarding factors that may influence both smoking transition behaviour and subsequent health outcomes. Age, sex, and ethnicity were included to account for demographic differences in smoking patterns, e-cigarette uptake, and baseline physical and mental health. Highest education attained, equivalised household income, employment status, and Government Office Region were incorporated as indicators of socioeconomic position and structural context, which are strongly associated with smoking prevalence, cessation success, access to healthcare, and health-related quality of life. Household net income was equivalised using the Organisation for Economic Co-operation and Development (OECD)-modified scale to account for household size and composition, adjusted for inflation using the Consumer Price Index to reflect real purchasing power over time, and subsequently log-transformed to reduce skewness and improve model interpretability (23). Long-standing illness was included to capture pre-existing health conditions that may influence both the likelihood of smoking transitions and subsequent health outcomes, helping to address confounding by indication. Annual GP visits were used as a proxy for healthcare utilisation and underlying health needs, while the number of children in the household was included to reflect household responsibilities and psychosocial factors that may affect smoking behaviour and stress-related health outcomes. Finally, lagged health outcomes (PCS, MCS, and EQ-5D-3L at baseline) were included to control for baseline health status, ensure appropriate temporal ordering, and reduce bias from reverse causation, particularly for mental health outcomes.

### Causal Framework: Directed Acyclic Graph (DAG)

A Directed Acyclic Graph (DAG) (*Figure 1 & 2*) was developed to formalise causal assumptions regarding the relationships between smoking transitions, health outcomes, and potential confounders (24, 25). The DAG was used to identify a minimal sufficient adjustment set required to block non-causal backdoor pathways between smoking transition status and physical health, mental health, and HRQoL outcomes.

**Figure 1.**
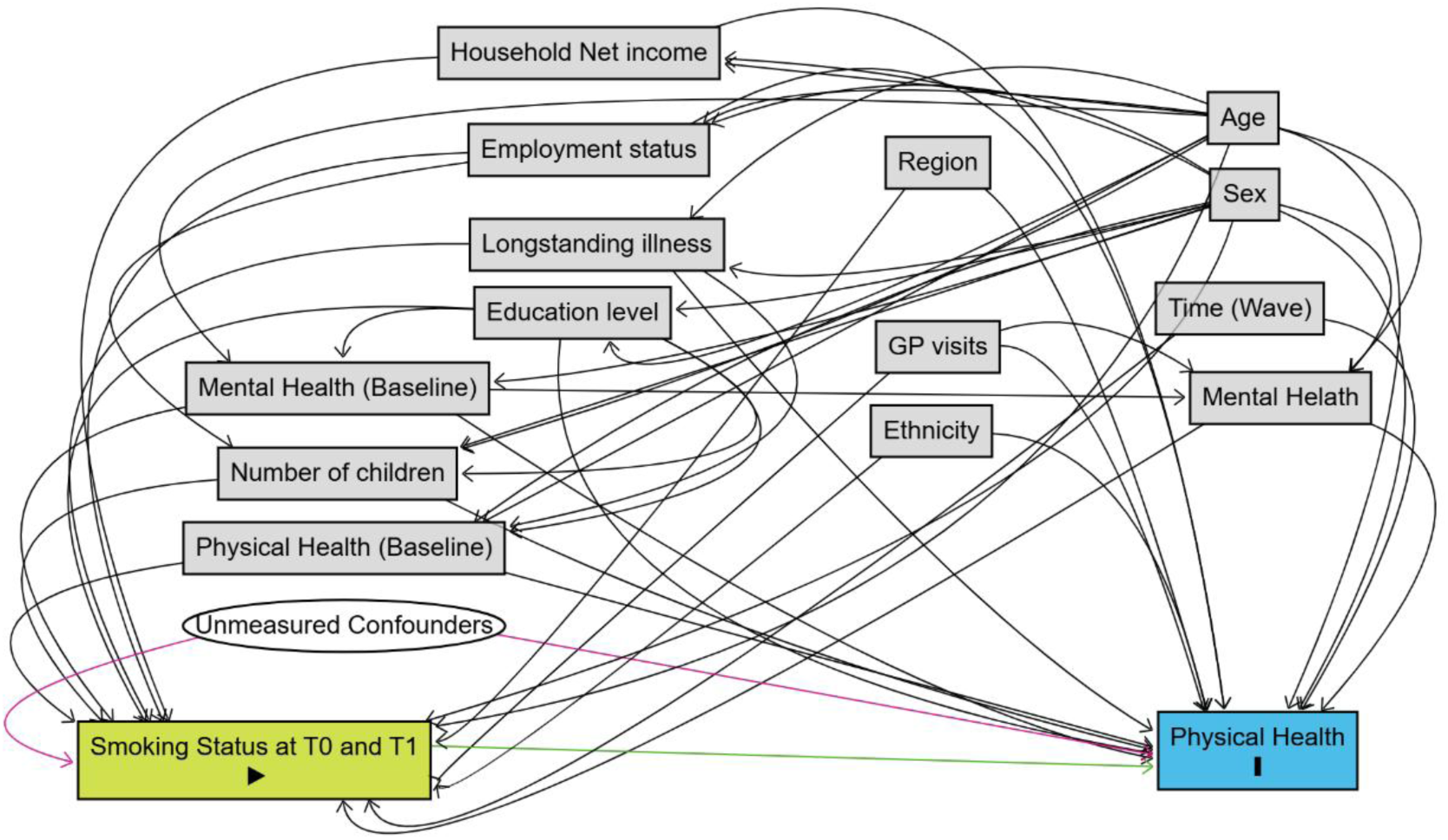
**Directed Acyclic Graph (DAG) for the Effect of Smoking Status at T0 and T1 on Physical Health**

**Figure 2.**
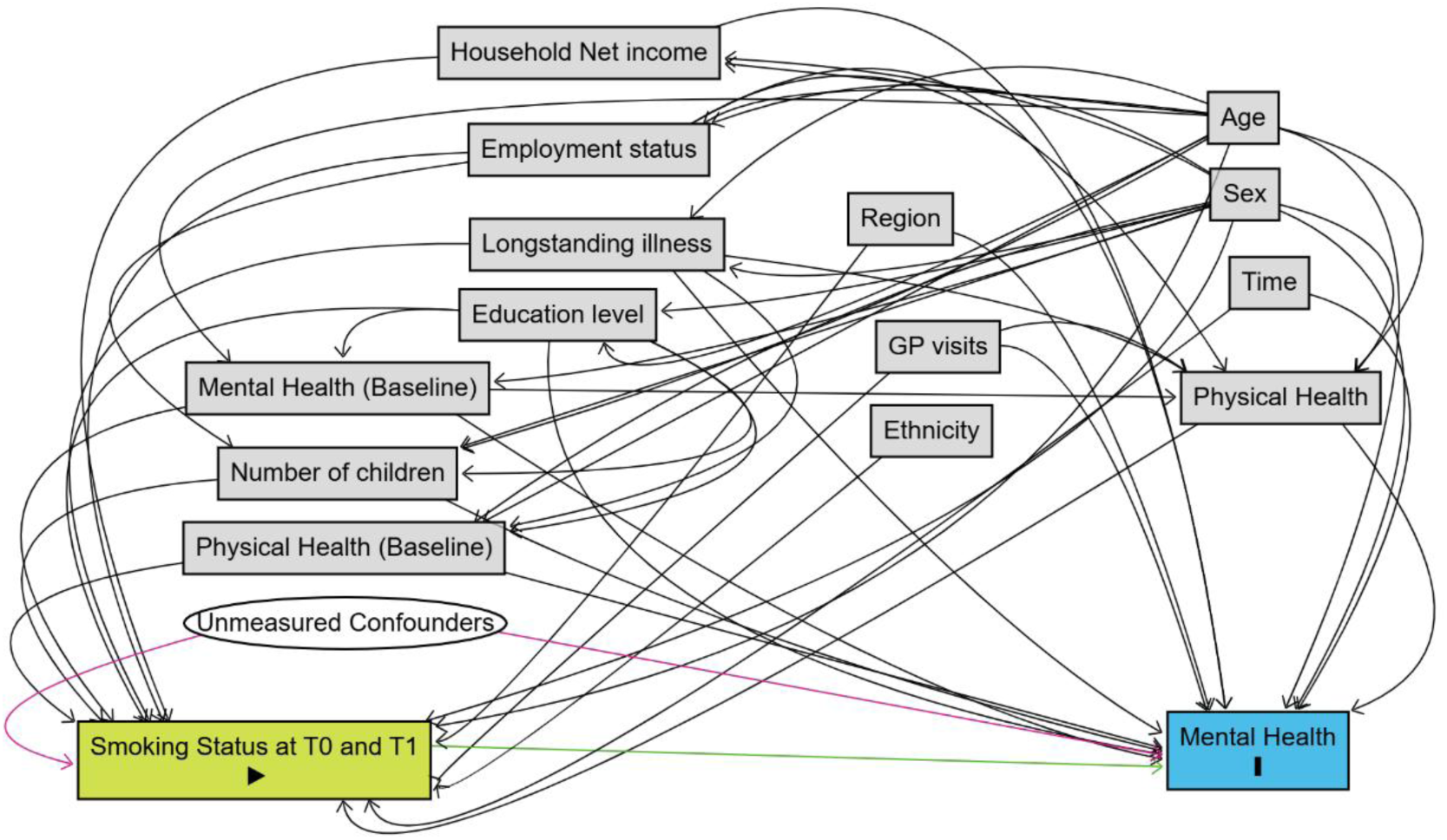
**Directed Acyclic Graph (DAG) for the Effect of Smoking Status at T0 and T1 on Mental Health**

### Statistical Analysis

A sequence of regression models was estimated to examine robustness across levels of adjustment:

- **Unadjusted OLS (unmatched)**: Bivariate associations in the full cohort.
- **Fully adjusted OLS (unmatched)**: Multivariable models adjusting for all baseline covariates.
- **Unadjusted OLS (matched)**: Comparisons within the propensity score–matched cohort.
- **Fully adjusted OLS (matched)**: The primary doubly robust model, combining propensity score matching with regression adjustment.
- **Fixed-effects models (matched)**: Sensitivity analyses controlling for time-invariant unobserved confounding.
- **Subgroup analyses**: Stratified analyses by age group, educational attainment, and income quintile to assess effect heterogeneity.

Propensity score matching was conducted using a 1:3 nearest-neighbour approach. Cluster-robust standard errors were applied in all models to account for repeated observations at the individual level (26) .

### Ethical Considerations

Ethical approval for the UKHLS was obtained by the original data collectors. This study involved secondary analysis of anonymised data, and no additional ethical approval was required.

## RESULTS

From 33,694 smokers identified across Waves 7-14 of the UK Household Longitudinal Study, 30,281 participants with complete demographic and exposure information were retained for eligibility screening. Restricting the sample to adults (≥16 years) who were exclusive traditional cigarette smokers at baseline yielded 24,189 eligible individuals. Requiring valid baseline and follow-up responses resulted in an unmatched analytical cohort of 18,011 participant-wave transitions (*Figure 3*).

**Figure 3.**
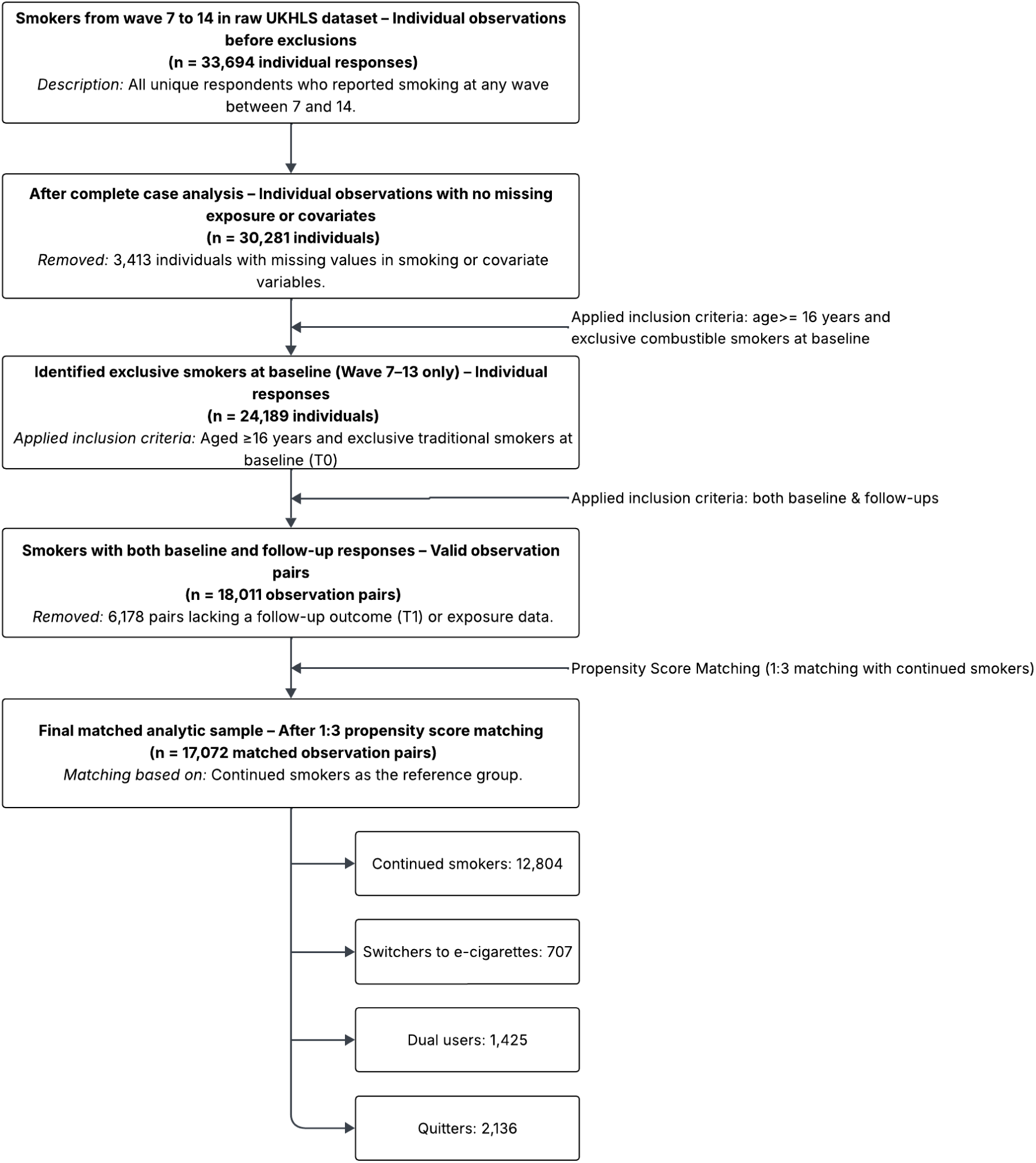
Flowchart of Study Sample Selection

At follow-up, participants were classified into four mutually exclusive smoking transition groups: continued smokers (n = 13,743; 76.3%), exclusive e-cigarette switchers (n = 707; 3.9%), dual users (n = 1,425; 7.9%), and quitters (n = 2,136; 11.9%). Thus, exclusive switching to e-cigarettes was relatively uncommon, while dual use represented a more frequent transitional pattern.

Baseline characteristics of the unmatched cohort, stratified by transition group, are presented in *Table 1*. Statistically significant differences were observed across groups for age, income, baseline physical health, sex, ethnicity, education, employment status, longstanding illness, healthcare utilisation, and region (all p < 0.05). These differences indicate substantial potential for confounding and motivate the use of causal inference methods.

**Table 1.** Baseline Characteristics of the Unmatched Cohort, Stratified by Smoking Transition Group.

| Characteristic | Level | Continued Smoker | Switcher | Dual User | Quitter | p value |
| --- | --- | --- | --- | --- | --- | --- |
| n |  | 13743 | 707 | 1425 | 2136 |  |
| Age (mean (SD)) |  | 48.28 (15.90) | 42.73<br>(14.51) | 44.62 (15.37) | 44.24<br>(16.82) | <0.001 |
| Log of Household Income (mean (SD)) |  | 7.16 (0.56) | 7.22<br>(0.62) | 7.18 (0.53) | 7.22<br>(0.69) | <0.001 |
| Physical Component summary at baseline (PCS/T<sub>0</sub>)<br>(mean (SD)) |  | 47.13 (11.95) | 49.05<br>(11.37) | 47.47 (11.95) | 49.26<br>(11.78) | <0.001 |
| Mental Component summary at baseline (MCS/T<sub>0</sub>)<br>(mean (SD)) |  | 45.53 (11.93) | 45.30<br>(12.02) | 45.20 (11.90) | 45.61<br>(11.89) | 0.709 |
| Physical Component summary at follow-up (PCS/T<sub>1</sub>)<br>(mean (SD)) |  | 46.68 (12.06) | 48.11<br>(11.76) | 46.92 (12.21) | 48.18<br>(11.88) | <0.001 |
| Mental Component summary at follow-up (MCS/ T<sub>1</sub>)<br>(mean (SD)) |  | 45.39 (12.02) | 45.61<br>(11.85) | 44.02 (11.95) | 45.74<br>(11.35) | <0.001 |
| Number of Kids |  | 0.52 (0.95) | 0.65<br>(1.04) | 0.57 (0.98) | 0.59<br>(0.95) | <0.001 |
| Sex (n (%)) | Female | 7352 (53.5) | 389<br>(55.0) | 713 (50.0) | 1168<br>(54.7) | 0.034 |
|  | Male | 6391 (46.5) | 318<br>(45.0) | 712 (50.0) | 968<br>(45.3) |  |
| Ethnicity (n (%)) | Asian | 743 (5.4) | 34 (4.8) | 66 (4.6) | 159<br>(7.4) | <0.001 |
|  | Black | 392 (2.9) | 15 (2.1) | 26 (1.8) | 86 (4.0) |  |
|  | Mixed | 366 (2.7) | 18 (2.5) | 46 (3.2) | 89 (4.2) |  |
|  | Other | 69 (0.5) | 2 (0.3) | 11 (0.8) | 9 (0.4) |  |
|  | White | 12173 (88.6) | 638<br>(90.2) | 1276 (89.5) | 1793<br>(83.9) |  |
| Highest Educational Qualification (n (%)) | A-level or<br>equivalent | 3179 (23.1) | 175<br>(24.8) | 329 (23.1) | 537<br>(25.1) | <0.001 |
|  | Degree or<br>higher | 3245 (23.6) | 204<br>(28.9) | 360 (25.3) | 734<br>(34.4) |  |
|  | GCSE or<br>equivalent | 3844 (28.0) | 201<br>(28.4) | 411 (28.8) | 489<br>(22.9) |  |
|  | No qualification | 1871 (13.6) | 62 (8.8) | 159 (11.2) | 201 (9.4) |  |
|  | Other qualification | 1604 (11.7) | 65 (9.2) | 166 (11.6) | 175 (8.2) |  |
| Employment Status (n (%)) | Economically inactive | 4980 (36.2) | 157 (22.2) | 423 (29.7) | 652 (30.5) | <0.001 |
|  | Employed | 7637 (55.6) | 507 (71.7) | 889 (62.4) | 1337 (62.6) |  |
|  | Unemployed | 1126 (8.2) | 43 (6.1) | 113 (7.9) | 147 (6.9) |  |
| Longterm Illness (n (%)) | No | 7968 (58.0) | 442 (62.5) | 830 (58.2) | 1342 (62.8) | <0.001 |
|  | Yes | 5775 (42.0) | 265 (37.5) | 595 (41.8) | 794 (37.2) |  |
| Annual GP visits (n (%)) | Zero | 3915 (28.5) | 205 (29.0) | 372 (26.1) | 565 (26.5) | 0.001 |
|  | One to five | 7608 (55.4) | 373 (52.8) | 779 (54.7) | 1251 (58.6) |  |
|  | Six to ten | 1127 (8.2) | 70 (9.9) | 133 (9.3) | 187 (8.8) |  |
|  | More than ten | 1093 (8.0) | 59 (8.3) | 141 (9.9) | 133 (6.2) |  |
| Regions (n (%)) | East Midland | 1016 (7.4) | 69 (9.8) | 97 (6.8) | 132 (6.2) | <0.001 |
|  | East of England | 1122 (8.2) | 62 (8.8) | 129 (9.1) | 187 (8.8) |  |
|  | London | 1186 (8.6) | 50 (7.1) | 89 (6.2) | 282 (13.2) |  |
|  | North East | 533 (3.9) | 40 (5.7) | 57 (4.0) | 78 (3.7) |  |
|  | North West | 1326 (9.6) | 75 (10.6) | 168 (11.8) | 197 (9.2) |  |
|  | Northern Ireland | 1069 (7.8) | 38 (5.4) | 87 (6.1) | 201 (9.4) |  |
|  | Scotland | 1421 (10.3) | 60 (8.5) | 143 (10.0) | 160 (7.5) |  |
|  | South East | 1422 (10.3) | 77 (10.9) | 158 (11.1) | 250 (11.7) |  |
|  | South West | 1138 (8.3) | 61 (8.6) | 132 (9.3) | 171 (8.0) |  |
|  | Wales | 1245 (9.1) | 63 (8.9) | 99 (6.9) | 149 (7.0) |  |
|  | West Midland | 1120 (8.1) | 45 (6.4) | 123 (8.6) | 155 (7.3) |  |
|  | Yorkshire and the Humber | 1145 (8.3) | 67 (9.5) | 143 (10.0) | 174 (8.1) |  |
| Wave (n(%)) | 7 | 3038 (22.1) | 213 (30.1) | 487 (34.2) | 472 (22.1) | <0.001 |
|  | 8 | 2318 (16.9) | 119 (16.8) | 217 (15.2) | 373 (17.5) |  |
|  | 9 | 2106 (15.3) | 87 (12.3) | 176 (12.4) | 287 (13.4) |  |
|  | 10 | 1904 (13.9) | 81 (11.5) | 144 (10.1) | 278 (13.0) |  |
|  | 11 | 1692 (12.3) | 42 (5.9) | 110 (7.7) | 316<br>(14.8) |  |
|  | 12 | 1482 (10.8) | 80 (11.3) | 149 (10.5) | 212<br>(9.9) |  |
|  | 13 | 1203 (8.8) | 85 (12.0) | 142 (10.0) | 198<br>(9.3) |  |

*Note: The p-value tests for any statistically significant difference in the distribution or mean of a characteristic across all four smoking transition groups. P-values are derived from ANOVA for continuous variables and Chi-square tests for categorical*

In unadjusted analyses comparing follow-up outcomes across the four groups, mean physical (PCS) and mental health (MCS) scores differed significantly (ANOVA p < 0.001 for both). Relative to continued smokers, switchers and quitters showed higher PCS and EQ-5D-3L scores, while dual users had lower MCS scores (*Table 2*).

**Table 2.**
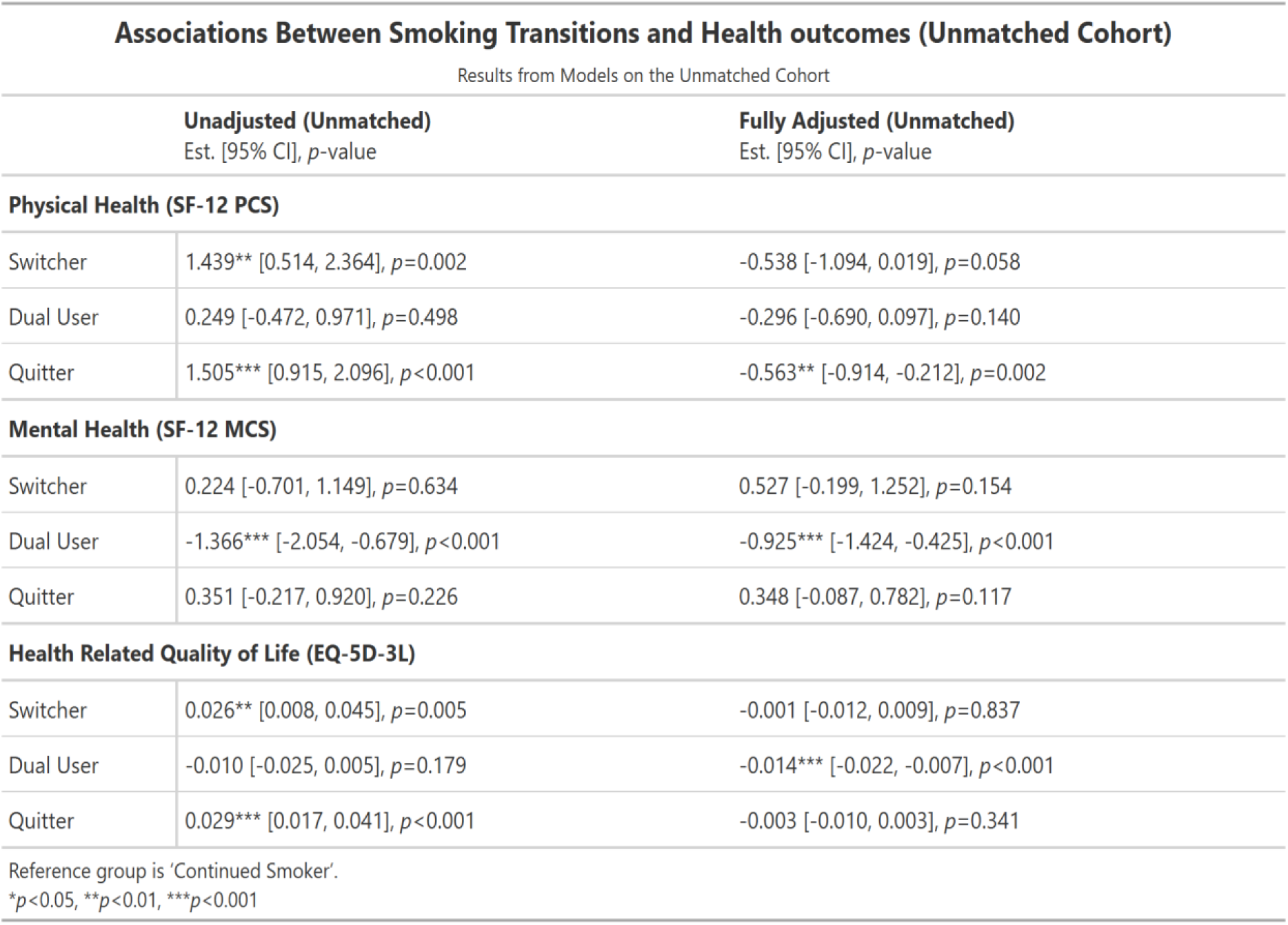
Unadjusted and Fully Adjusted Associations Between Smoking Transitions and Health Outcomes in the Unmatched Cohort.

| **Associations Between Smoking Transitions and Health outcomes (Unmatched Cohort)** |  |  |
| --- | --- | --- |
| Results from Models on the Unmatched Cohort |  |  |
| | **Unadjusted (Unmatched)**<br>Est. [95% CI], $p$-value | **Fully Adjusted (Unmatched)**<br>Est. [95% CI], $p$-value |
| **Physical Health (SF-12 PCS)** |  |  |
| Switcher | 1.439** [0.514, 2.364], $p=0.002$ | -0.538 [-1.094, 0.019], $p=0.058$ |
| Dual User | 0.249 [-0.472, 0.971], $p=0.498$ | -0.296 [-0.690, 0.097], $p=0.140$ |
| Quitter | 1.505*** [0.915, 2.096], $p<0.001$ | -0.563** [-0.914, -0.212], $p=0.002$ |
| **Mental Health (SF-12 MCS)** |  |  |
| Switcher | 0.224 [-0.701, 1.149], $p=0.634$ | 0.527 [-0.199, 1.252], $p=0.154$ |
| Dual User | -1.366*** [-2.054, -0.679], $p<0.001$ | -0.925*** [-1.424, -0.425], $p<0.001$ |
| Quitter | 0.351 [-0.217, 0.920], $p=0.226$ | 0.348 [-0.087, 0.782], $p=0.117$ |
| **Health Related Quality of Life (EQ-5D-3L)** |  |  |
| Switcher | 0.026** [0.008, 0.045], $p=0.005$ | -0.001 [-0.012, 0.009], $p=0.837$ |
| Dual User | -0.010 [-0.025, 0.005], $p=0.179$ | -0.014*** [-0.022, -0.007], $p<0.001$ |
| Quitter | 0.029*** [0.017, 0.041], $p<0.001$ | -0.003 [-0.010, 0.003], $p=0.341$ |
| Reference group is 'Continued Smoker'. |  |  |
| *$p<0.05$, **$p<0.01$, ***$p<0.001$ | | |

After adjustment for baseline covariates, these apparent physical health advantages for switchers and quitters were no longer observed. However, dual use remained associated with poorer mental health compared with continued smoking, and quitters showed lower short-term physical health scores. These changes in estimates suggest substantial confounding in unadjusted models.

Following 1:3 propensity score matching, good covariate balance was achieved for all pairwise comparisons with continued smokers, with standardised mean differences below 0.1 (*Figures 4-6*).

**Figure 4.**
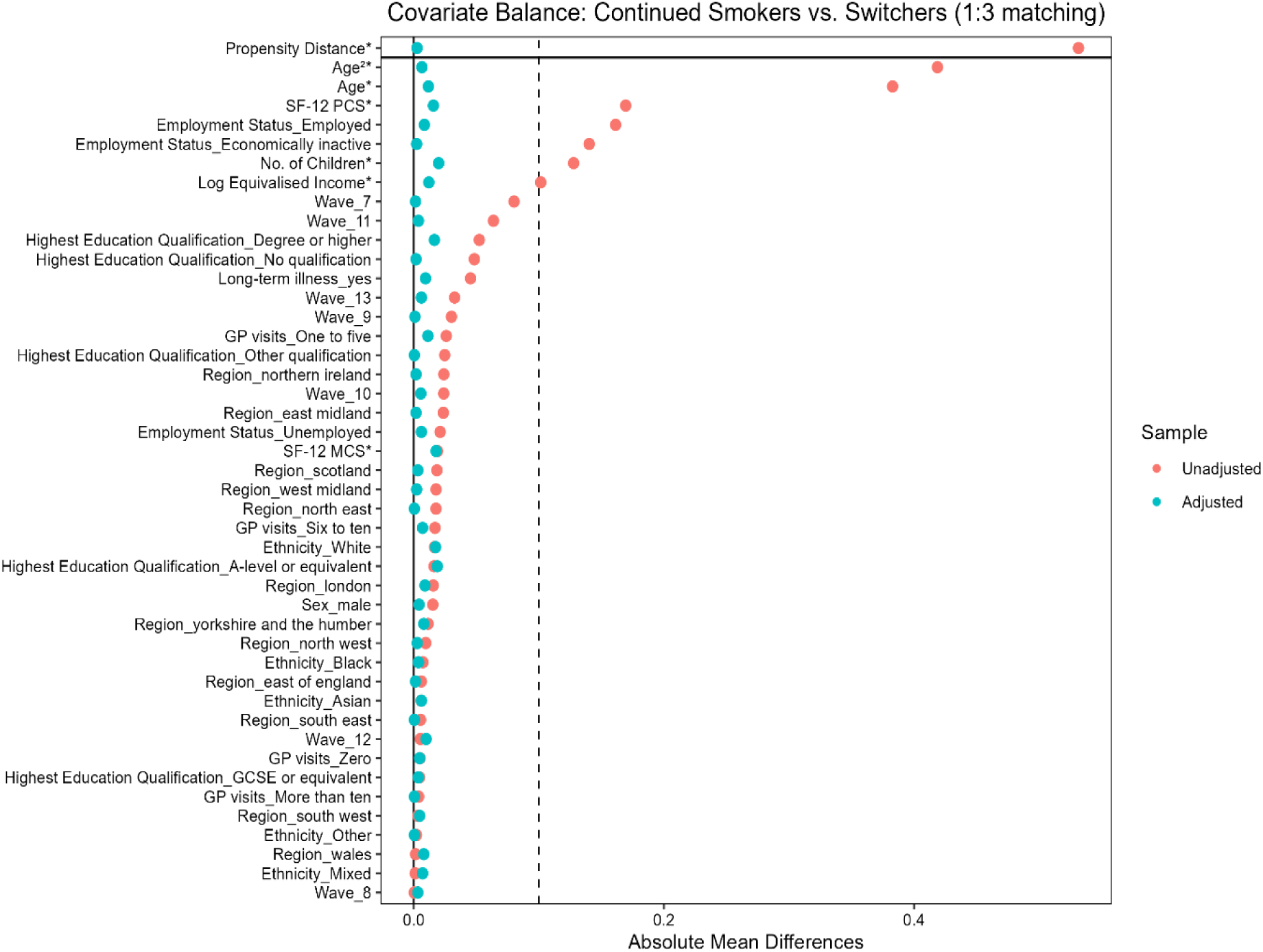
Covariate Balance for Continued Smokers vs Switchers (1:3 Matching)

**Figure 5.**
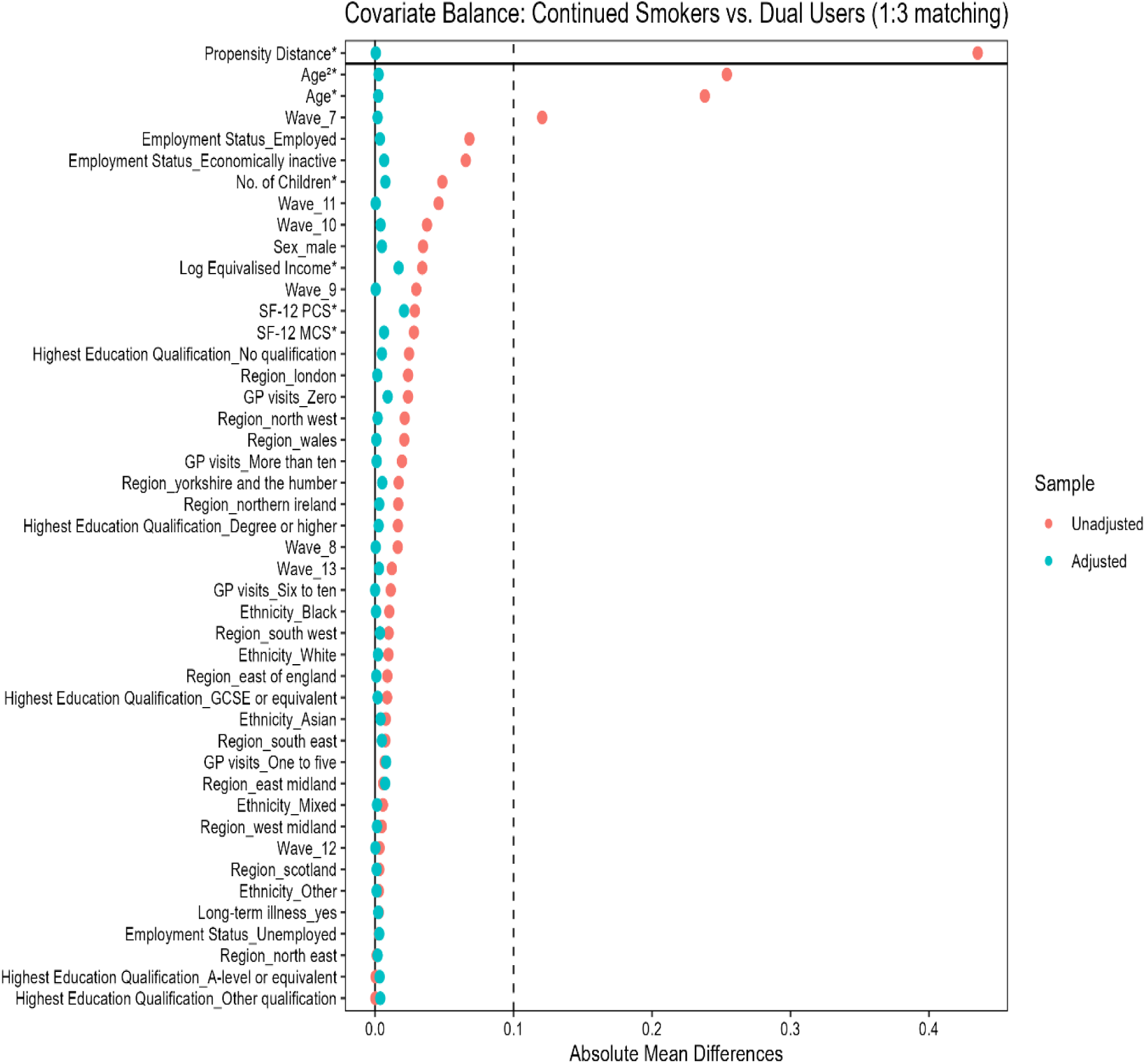
Covariate Balance for Continued Smokers vs Dual Users (1:3 Matching)

**Figure 6.**
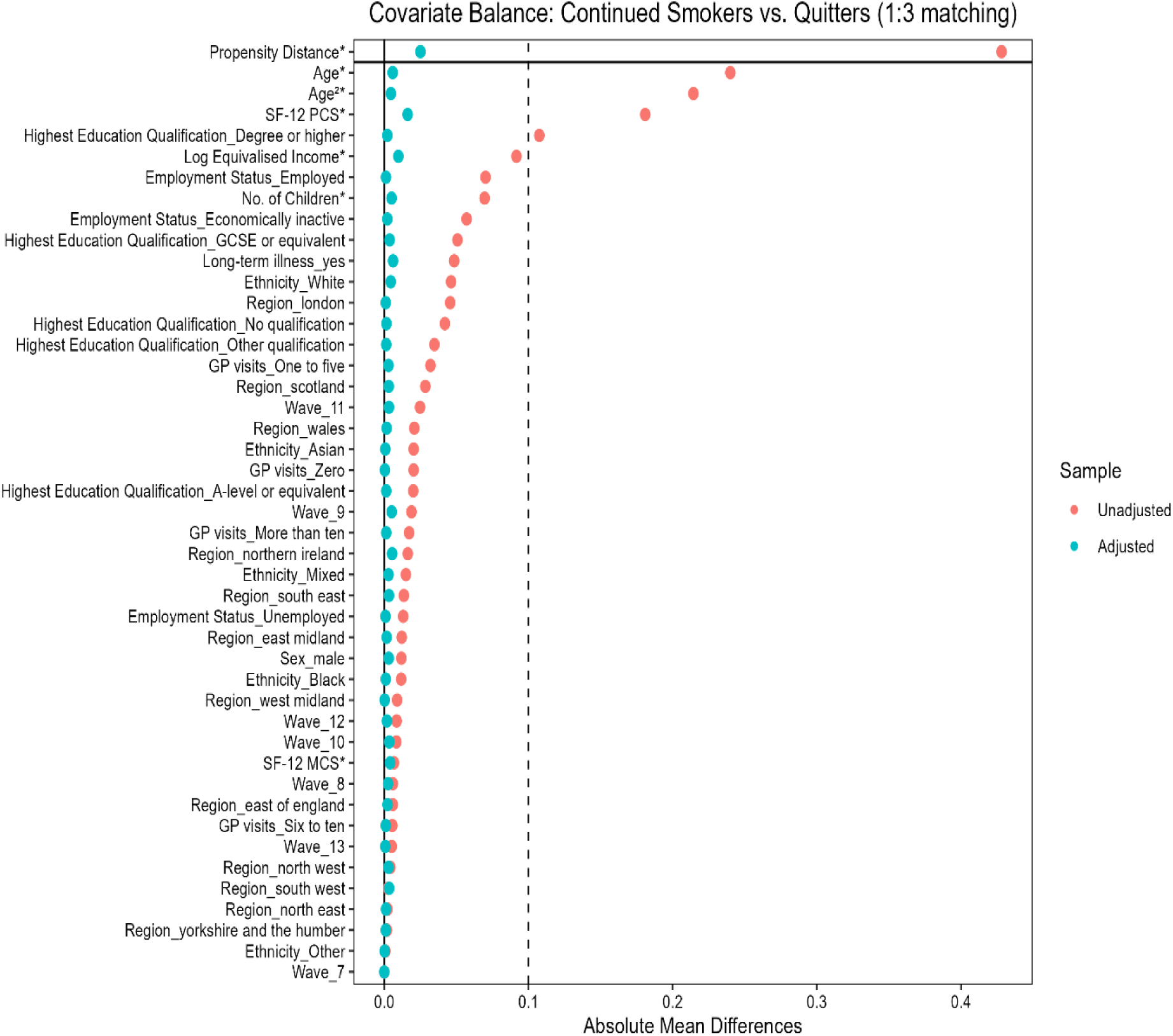
Covariate Balance for Continued Smokers vs Quitters (1:3 Matching)

In the doubly robust models (*Table 3*), several consistent patterns emerged:

- Physical health (PCS): Compared with continued smokers, both switchers (β = –0.670, p = 0.035) and quitters (β = –0.486, p = 0.010) had lower short-term PCS scores.
- Mental health (MCS): Switchers experienced improved mental health relative to continued smokers (β = 1.042, p = 0.012), whereas dual users had significantly poorer mental health (β = –1.023, p < 0.001).
- Health-related quality of life (EQ-5D-3L): Dual use was associated with lower HRQoL (β = –0.016, p < 0.001), while estimates for switchers and quitters were not statistically different from continued smokers

**Table 3.** Unadjusted and Fully Adjusted Associations Between Smoking Transitions and Health Outcomes in the Matched Cohort.

| Associations Between Smoking Transitions and Health outcomes (1:3 PS matched Cohort) |  |  |
| --- | --- | --- |
| Results from Models on the Matched Cohort |  |  |
|  | Unadjusted Model<br>Est. [95% CI], *p*-value | Doubly Robust Model<br>Est. [95% CI], *p*-value |
| **Physical Health (SF-12 PCS)** |  |  |
| Switcher | -0.479 [-1.485, 0.527], *p*=0.350 | -0.670* [-1.292, -0.048], *p*=0.035 |
| Dual User | -0.218 [-0.995, 0.558], *p*=0.582 | -0.391 [-0.825, 0.043], *p*=0.077 |
| Quitter | -0.330 [-0.938, 0.279], *p*=0.288 | -0.486** [-0.853, -0.119], *p*=0.010 |
| **Mental Health (SF-12 MCS)** |  |  |
| Switcher | 1.168* [0.129, 2.208], *p*=0.028 | 1.042* [0.229, 1.855], *p*=0.012 |
| Dual User | -1.038** [-1.790, -0.286], *p*=0.007 | -1.023*** [-1.574, -0.472], *p*<0.001 |
| Quitter | 0.393 [-0.210, 0.997], *p*=0.202 | 0.346 [-0.116, 0.807], *p*=0.142 |
| **Health Related Quality of Life (EQ-5D-3L)** |  |  |
| Switcher | 0.006 [-0.014, 0.026], *p*=0.548 | 0.003 [-0.009, 0.015], *p*=0.639 |
| Dual User | -0.014 [-0.030, 0.003], *p*=0.100 | -0.016*** [-0.025, -0.008], *p*<0.001 |
| Quitter | -0.000 [-0.012, 0.012], *p*=0.964 | -0.002 [-0.009, 0.005], *p*=0.510 |
| Note: Reference group is 'Continued Smoker'. |  |  |
| * *p* <0.05, ** *p* <0.01, *** *p* <0.001 |  |  |

Sensitivity analyses yielded results consistent with the primary findings. Estimates from fixed-effects models, which additionally control for time-invariant individual characteristics, showed continued negative associations for dual users in mental health and HRQoL and for quitters in physical health (*Table 4*).

**Table 4.**
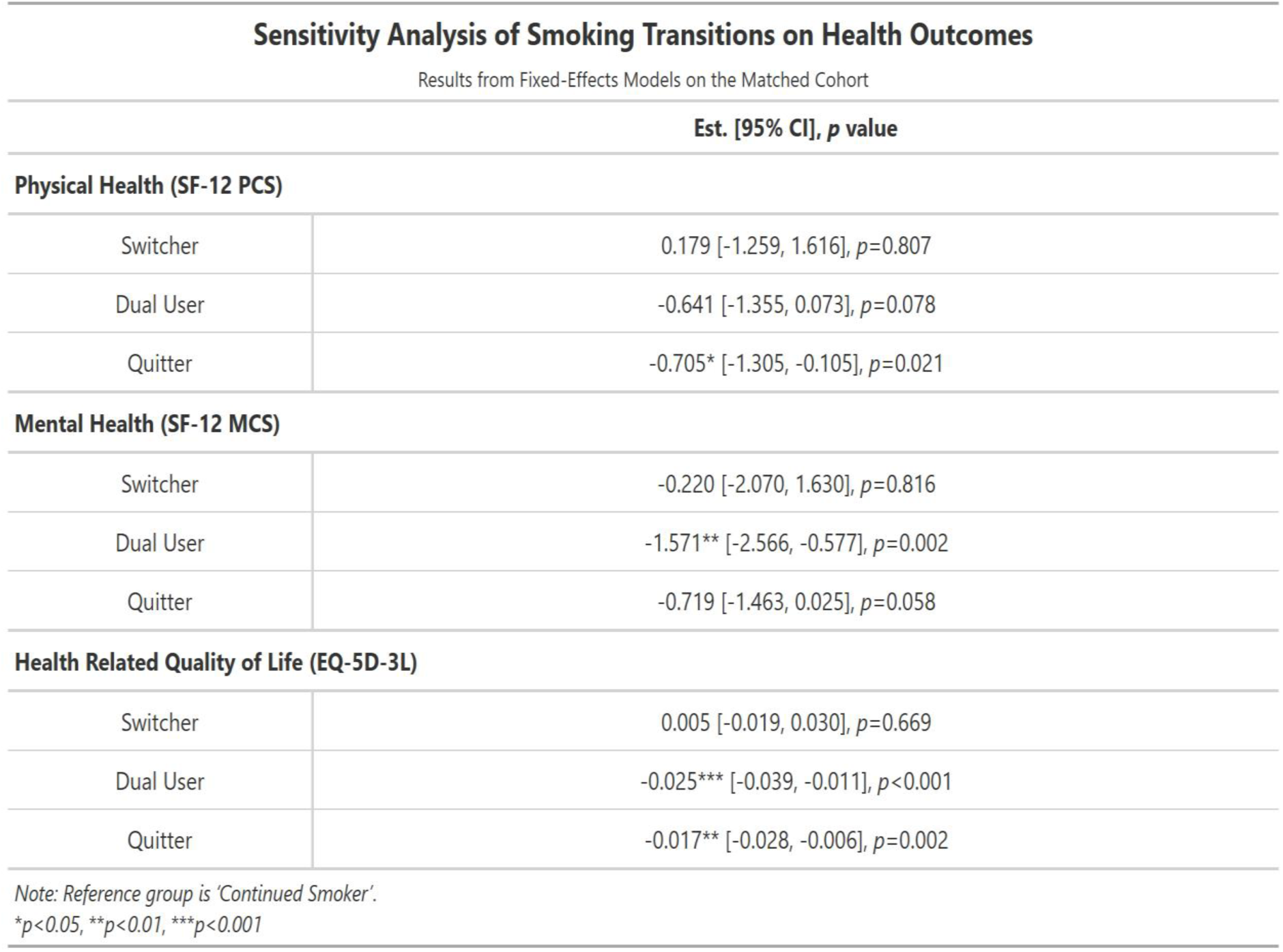
Sensitivity Analysis of Smoking Transitions on Health Outcomes using Fixed-Effects Model.

Using a 1:1 matching ratio produced similar effect estimates but with reduced precision due to smaller sample size. Subgroup analyses revealed heterogeneity by age, education, and income. Among younger adults (16–24 years), quitting was associated with improved mental health, whereas among older adults (≥50 years), switching and dual use were associated with poorer physical or mental health outcomes. Educational gradients were also evident: among individuals with no formal qualifications, switching was associated with improved mental health but poorer physical health, while dual use was consistently associated with adverse outcomes among those with higher educational attainment. Income-stratified analyses showed less consistent patterns, though negative physical health associations were observed in some middle-income groups.

Figure 7-9 summarises estimates across all analytical models. Apparent benefits of switching and quitting observed in unadjusted analyses attenuated or reversed following confounding adjustment, particularly for physical health. In contrast, the negative association between dual use and mental health remained consistent across models, reinforcing the robustness of this finding.

**Figure 7.**
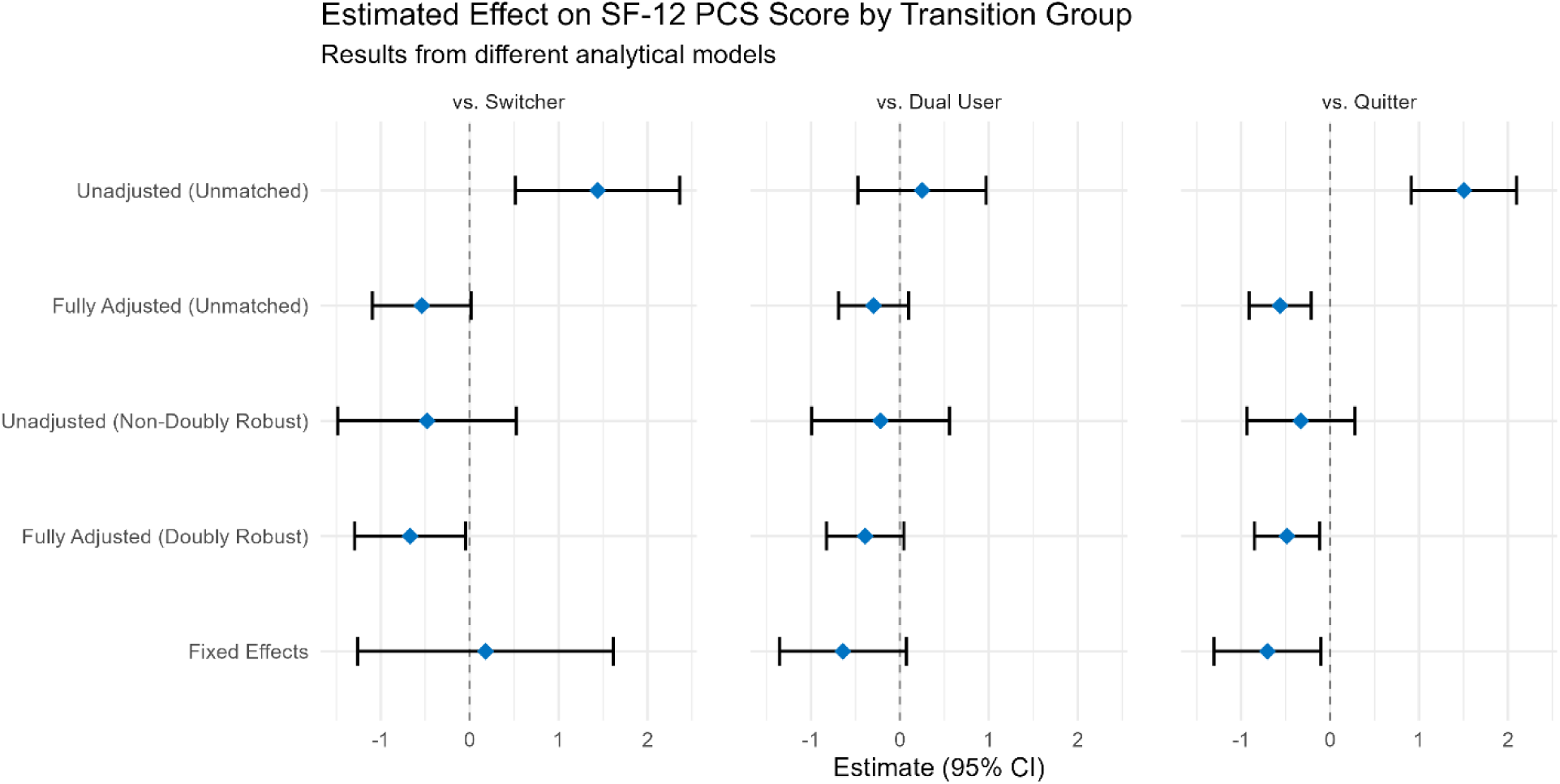
Forest Plot Comparing the Estimated Effect on SF-12 PCS Score Across Five Analytical Models.

**Figure 8.**
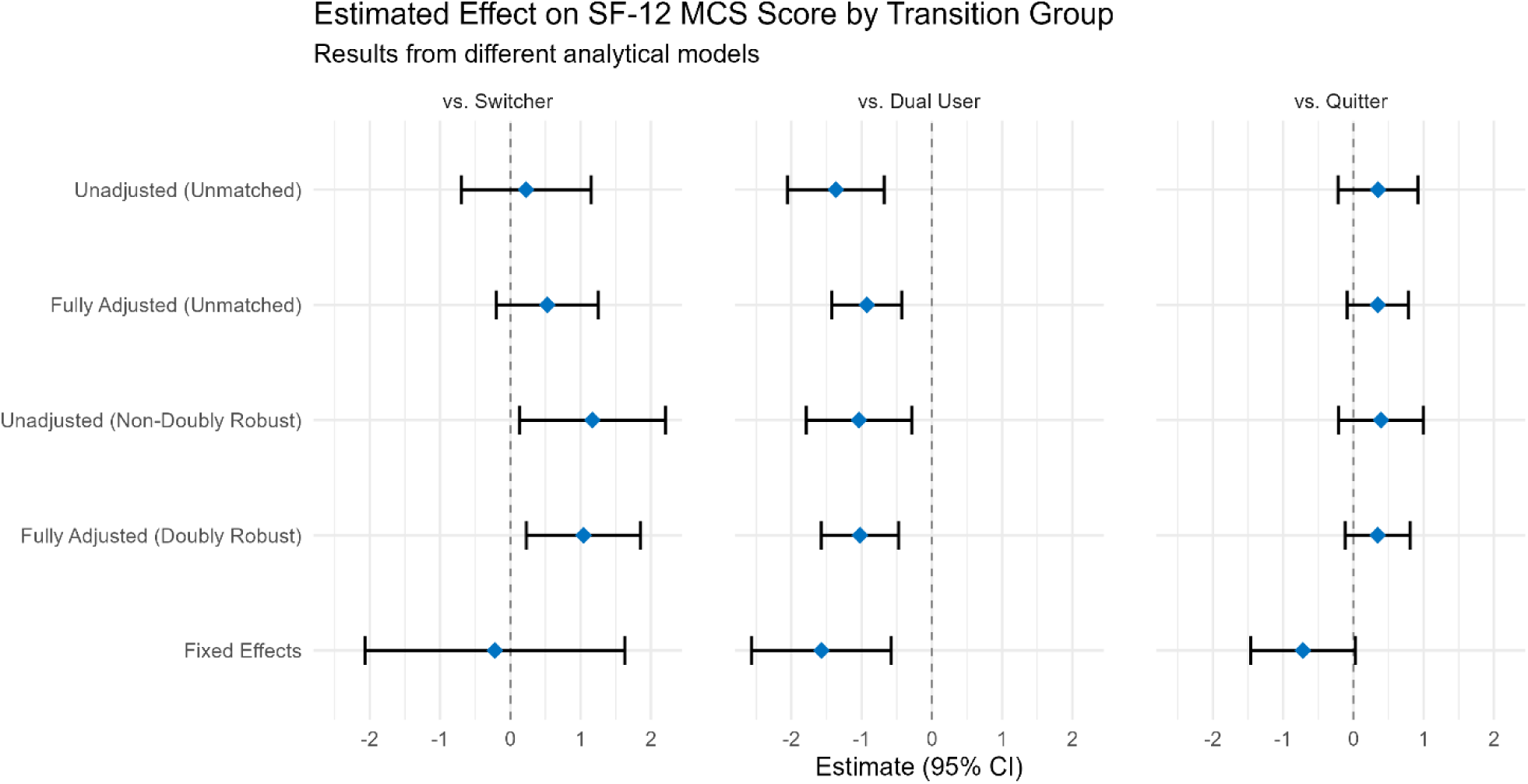
Forest Plot Comparing the Estimated Effect on SF-12 MCS Score Across Five Analytical Models.

**Figure 9.**
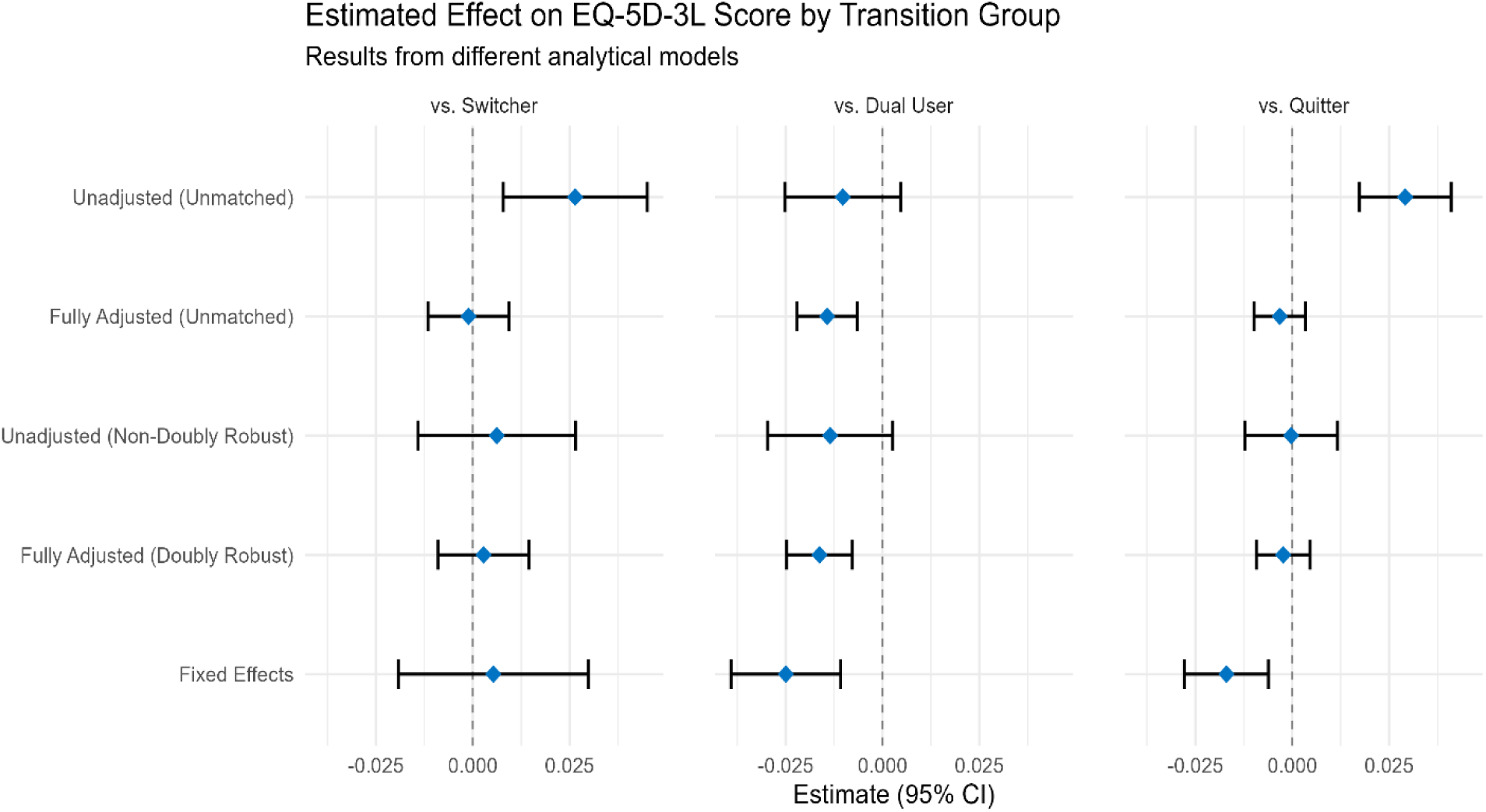
Forest Plot Comparing the Estimated Effect on EQ5D Score Across Five Analytical Models

## DISCUSSION

In this longitudinal analysis of UK smokers, exclusive switching to e-cigarettes was associated with improved short-term mental health compared with continued smoking, whereas dual use was consistently associated with poorer mental health and lower health-related quality of life (HRQoL). No short-term improvements in physical health were observed for any smoking transition group. Importantly, the apparent physical health “benefits” seen in unadjusted analyses disappeared after accounting for baseline differences, highlighting the presence of confounding by indication. These findings contrast sharply with results from simplistic models and align with a growing body of evidence suggesting that partial substitution through dual use offers limited benefit, while complete switching may confer short-term psychological advantages within a harm-reduction framework.

The low incidence of exclusive switching to e-cigarettes observed in this study (3.9%) is consistent with findings from the Scottish Health Survey (27) and other international studies, which show that although smoking prevalence is declining, dual use remains a common transitional state (6). This suggests that many smokers supplement rather than replace cigarette use with e-cigarettes.

With respect to physical health, the absence of short-term improvement is consistent with biological understanding of smoking-related harm (1, 2). Physical health damage reflects cumulative exposure over decades, and recovery involves slow cellular and physiological repair processes that are unlikely to manifest as detectable changes in broad self-reported measures over a single year. This contrasts with unadjusted findings in some studies and in the naïve models here, which likely reflect confounding by indication rather than true health effects (12).

Mental health outcomes showed clearer differentiation between transition groups. The improvement observed among exclusive switchers supports the potential psychological dimension of harm reduction, while declines among quitters likely reflect short-term withdrawal, stress, or loss of a coping mechanism (28). Age and socioeconomic subgroup patterns mirror established inequalities, with younger individuals showing more favourable mental health outcomes and disadvantaged groups experiencing greater challenges, reinforcing evidence that smoking is closely intertwined with social stress and coping (29–31).

HRQoL results confirm its multidimensional nature. Improvements in mental health among some groups were offset by a lack of corresponding physical health gains, resulting in largely neutral overall changes (32, 33). This highlights the importance of interpreting HRQoL as a balance of multiple health domains rather than a single unified outcome.

This study has several strengths. First, it uses data from the UK Household Longitudinal Study, a large and broadly nationally representative dataset, enhancing the relevance of findings for the UK population. Second, the longitudinal design allows repeated observations within individuals, enabling the examination of within-person smoking transitions rather than static smoking status. Third, the application of a target trial emulation framework with propensity score matching and regression adjustment strengthens causal interpretation relative to conventional observational approaches. Finally, the consistency of findings across multiple analytical strategies, including matched, doubly robust, and fixed-effects models, supports the robustness of the results.

Several limitations should be acknowledged. The short follow-up period limits the ability to detect physical health recovery, which is known to occur gradually over longer time horizons. Health outcomes were self-reported, introducing potential measurement error, particularly during periods of withdrawal or psychological distress. Residual confounding from time-varying factors, such as nicotine dependence severity or psychiatric comorbidities, may remain despite adjustment. Reverse causality is also possible, whereby deteriorating health prompts quitting rather than resulting from it. Finally, the use of complete case analysis may reduce generalisability if individuals with missing follow-up data differ systematically from those retained in the analysis.

Future research should prioritise longer follow-up periods to capture slow physical health recovery and long-term outcomes. Combining self-reported measures with objective data, such as biomarkers or linked medical records, would improve measurement of physical health changes. Addressing missing data through multiple imputation could enhance representativeness and statistical power. Finally, explicitly modelling time-varying confounding and health-triggered behaviour change will be essential to further reduce bias and strengthen causal inference in studies of smoking transitions.

## CONCLUSION

In this UK longitudinal study, exclusive switching from cigarettes to e-cigarettes was associated with improved short-term mental health, while dual use was consistently linked to poorer mental health and reduced quality of life. No short-term physical health benefits were observed, underscoring the slow biological recovery from smoking-related harm. These findings highlight the importance of distinguishing complete switching from dual use in harm-reduction policy and smoking cessation support.

## Data Availability

All data produced are available online at https://datacatalogue.ukdataservice.ac.uk/studies/study/7629#details

https://github.com/shkjsm/shkjsm-smoking-transitions-health-UKHLS

